# Sexual Initiation and the Timing of First Union among women in Uganda

**DOI:** 10.1101/2025.01.31.25321478

**Authors:** Charles Lwanga, Dick Nsimbe, Ishmael kalule-Sabiti, Kudzaishe Mangombe, Jude Otim

## Abstract

The aim of this paper was to examine the socioeconomic correlates of the length of the gap between onset of sex initiation and the age at first union, and also estimate the waiting time to first union after sexual debut. Self-reported data from the individual record file were extracted from the 2016 Uganda Demographic and Health Survey (UDHS), comprising of a subsample of 4293 women aged 15-25. Chi-square tests and the logistic regression of multivariable event history analysis were used to investigate the relationship between socioeconomic factors and the length of the gap. Descriptive findings show that the average and mean waiting time in years is about 5.6 and 5.5 respectively. Age at sexual initiation and current age of the woman were the only significant factors. Women who had their sexual debut before age 15 would take longer to transit to first union and this is perhaps attributed to serial monogamy, and having multiple and concurrent partners which leads to a failure in subsequent union formation. Women aged 20-25 had a disproportionally lower risk of transiting to first union, which is attributed to social development aspects, diverging from earlier thinking that a woman is supposed to either be one’s wife or one’s mother. In conclusion, efforts to address what might influence the length of the gap between sexual union and first union need to take into account the age at sexual debut and the current age of the women.

## Introduction

Sexual behaviours in the interval between sexual debut and age at first union has been associated with immediate and longer-term demographic, social, and reproductive health consequences for the individual and the society (Batyra et al., 2021; Bongaarts, 2007; Zaba et al., 2009). This is because a woman who has a longer interval between age at first sex and age at first union is expected to have several sexual partners over the lifetime, which carries health and social consequences. In the context of HIV/AIDS, monitoring the trend in age at sexual debut and first union, has increasingly become important since interventions usually target young women (Chemhaka & Simelane, 2024; Ferede et al., 2023; Zaba et al., 2004). In fertility studies, early age at sexual debut and first marriage has been associated with a longer exposure to the risk of pregnancy, high fertility and high population growth rates (Bongaarts & Potter, 1983; Girmatsion et al., 2024).

Early age at childbearing has been investigated in relation to maternal morbidity, with some researchers suggesting that marrying early increases the risk of maternal mortality and morbidity (Melah et al., 2007; Tebeu et al., 2012); and others arguing that although late age at marriage is appropriate for women who pursue higher education, marrying late might lead to increased risk of HIV/AIDS and premarital births (Bongaarts, 2007). Using data from the 2003 Demographic and Health Surveys for Kenya and Ghana, Bongaarts (ibid) further found HIV prevalence to be associated with the length of the interval between first sexual intercourse and first union.

In Africa, research on the factors associated with the interval between age at first sex and age at first union and the implication such factors have for the reproductive health of women is limited. Existing studies focused on the influence age at sex initiation and age at first marriage have on women’s sexual behaviour (Zaba et al., 2009); on determinants of age at first marriage (Ayiga & Rampagane, 2013; Gurmu & Mace, 2013; McGrath et al., 2009); on the description of recent trends in age at first sex (Zaba et al., 2004); on age at first sex and HIV prevention strategies (Bakilana, 2005); and on the age at first marriage and HIV infection (Adair, 2007). In demographic studies however, least explored are the factors associated with length of the interval between age at sex initiation and the age at which individual form a first union and the reproductive health implication such factors may have for women later in life. In addition, little is known on the timing of first union after sex initiation.

In Uganda, data from the 2016 Uganda Demographic and Health Survey showed that the median age at first marriage is 17.9 years and age at first sex is 16.8 years, resulting in a 1.1-year gap between age at sexual debut and age at first marriage. Given that age at sexual debut and first union is not the same, this gap has implications for reproductive health for young women in future. First, it is a high-risk period involving sexual experimentation; second, relationships formed during this interval are usually transitory; third, STIs contracted during this period may pose a health risk to the future partner regardless of the faithfulness status later in union; and fourth, sexual behaviour patterns during this interval may shape the behaviour of young women in future. Drawing on these implications, the objective of this paper is twofold. First, to explore the sociodemographic factors associated with the length of the gap between onset of sex initiation and the age at first union later in life; and second, to estimate the waiting time to first union after sexual debut. The paper also discusses the implications of the results for reproductive health of young women in Uganda.

## Data and Methods

This paper used data from the 2016 Uganda Demographic and Health Survey (UDHS). Demographic and Health Surveys (DHS) are currently part of the worldwide survey programmes, which are a source of nationally representative estimates on individual and household level sociodemographic, health status and health care data in Uganda. UDHS collects information in the area of reproduction, marriage and sexual activity, maternal and child heath, mortality, fertility, family planning and nutrition. Following a two-stage sampling procedure, in which the first stage comprised of selecting enumeration areas (EAs) and in the second stage, households were selected; UDHS collected information from 8674 women aged 15-49 using a set of questionnaires consistent with the MEASURE DHS model questionnaire. This model ensures standardization, quality control procedures and common features across countries. In all households selected, all women of reproductive aged15-49 years were eligible for interview (UBOS & ICF International Inc, 2016).

### Variables

The dependent variable is the time to first union following sexual debut. The time was computed in years from questions on marriage and sexual activity: How old were you when you had sexual intercourse for the very first time? And how old were you when you first started living with a man? Based on each woman’s response, the time to first union was computed by subtracting the age of first sexual intercourse from the age of first union. A woman respondent was right censored if, by the time of the survey, she had not entered into first union. The analytical sample consisted of women who were aged between15 and 25 years inclusive. Further cleaning of the 2016 UDHS data was conducted and the analytical sample consisted of 4293 women (49.5%). The choice of age range (15 to 25 years) is based on three reasons: First, to minimize the effects of time; second, this age range is also fairly homogeneous with regard to cohort analysis; and third, by age 25, majority of the women would have finished schooling and entered into first union. The independent variables are sociodemographic in nature. They include factors which directly influence the time to first union; factors which indirectly affect the time to first union; and third category are the control variables. The direct factors were number of sexual partners in life time, having a child before union; indirect factors included are wealth quintile of the natal household, region of residence and place of residence. The control variables included are current age and early age at first sex.

### Statistical analysis

Survival analysis methods including lifetable (LT), Kaplan Meier (KM) survival curves and the logistic regression of multivariate event history analysis, namely the discrete-time to event model were used in data analysis to estimate the mean time from sexual debut to first union; and also identify the risk factors associated with the timing of first union given sexual debut. The lifetable method was used to estimate the proportion of women transiting to first union at the end of each interval after sex initiation (Preston et al., 2001). A generalised Wilcoxon test fixed at p<0.05 was used to test for the difference between survival functions of different groups. The test is non-parametric and was considered appropriate because some of the variables were not evenly distributed. To estimate the mean time to first union following sexual debut, the Kaplan Meier (KM) survival curve was used at bivariate level; and the significant difference in the meantime resulting from differences in covariates was tested using the Log-Rank test set at 5%. After changing the data from being person oriented to person-period, we fitted a discrete-time logit model. This is a binary response model to “*y*_*ti*_” (as shown in equation 1), to identify the risk factors associated with the timing of first union. The risk factors are usually estimated as the ratio of women entering first union at the end of the interval to those who had attained sexual debut. The fitted model was assessed using the link-test to examine whether the explanatory variables were specified correctly and also test for the goodness-of-fit (Cleves et al., 2010; Hilbe, unpublished; Kohler & Kreuter, 2012). The Link-test uses the hat and _hat-squared statistic and when the model describes the data correctly and is appropriate, the hat-squared should not be significant (_hat-squared, p>0.05). Before fitting the model, a multi-collinearity test among explanatory variables (results not presented) was conducted.

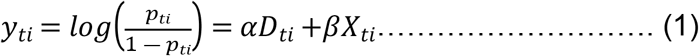

## Results

The mean age at sexual debut for this study was estimated to be 16 years and that of first union 18 years. Table 1 & Fig. 1 present women who have ever had sexual debut and entered first union by some background characteristics, mean length of the interval to first union by characteristics of interest using the Kaplan Meier estimates; and the Log Rank to test for the significance of the differences. The table shows that a woman’s child status, number of sexual partners, wealth index, region of residence, type of residence, whether age at sexual debut is less than 15 years, and woman’s current age were found to be strongly associated with the length of the interval (or the gap) between sexual debut and age at first union. The mean length of the gap was estimated to be about 5.6 years (see Table 1) and the median 5.5 years. Additionally, Fig.1 show that the transition to first union followed a few years after sexual debut.

**Table 1.** Proportion of women, who attained sexual debut by background characteristics, mean and median length of the gap, and log Rank chi^2^, Uganda.

| Covariates | N | Percentage distribution | Mean length of the gap (years) | Median length of the gap (Years) | χ<sup>2</sup>(chi<sup>2</sup>) |
| --- | --- | --- | --- | --- | --- |
| **Factors directly associated with the gap** |  |  |  |  |  |
| **Child status** |  |  |  |  |  |
| Never had a child | 600 | 77.12 | 6.3 | 6.0 | 78.21<br>(p=0.000) |
| Had a child in the gap before first union | 178 | 22.88 | 8.4 | 8.0 |  |
| **Number of sexual partners** |  |  |  |  |  |
| One partner | 1914 | 44.58 | 5.2 | 5.0 | 35.79<br>(p=0.000) |
| More than one partner | 2379 | 55.42 | 5.6 | 5.0 |  |
| **Factors indirectly associated with the gap** |  |  |  |  |  |
| **Wealth quintile** |  |  |  |  |  |
| Poorest | 1067 | 24.85 | 4.9 | 5.0 | 217.68<br>(p=0.000) |
| Poorer | 1000 | 23.29 | 5.1 | 5.0 |  |
| Middle | 769 | 17.91 | 5.4 | 5.0 |  |
| Richer | 738 | 17.19 | 5.2 | 6.0 |  |
| Richest | 719 | 16.75 | 6.4 | 6.0 |  |
| **Region** |  |  |  |  |  |
| Kampala | 230 | 5.36 | 6.5 | 7.0 | 79.94<br>(p=0.000) |
| Central | 694 | 16.17 | 5.7 | 6.0 |  |
| Eastern | 1228 | 28.60 | 5.3 | 5.0 |  |
| Northern | 1076 | 25.06 | 5.2 | 5.0 |  |
| Western | 1065 | 24.81 | 5.5 | 5.0 |  |
| **Type of residence** |  |  |  |  |  |
| Urban | 862 | 20.08 | 6.1 | 6.0 | 93.08<br>(p=0.000) |
| Rural | 3431 | 79.92 | 5.3 | 5.0 |  |
| **Controls Factors** |  |  |  |  |  |
| **Age at sexual debut <15 years** |  |  |  |  |  |
| Yes | 713 | 16.61 | 3.8 | 4.0 | 317.24<br>(p=0.000) |
| No | 3580 | 83.39 | 5.8 | 6.0 |  |
| **Age** |  |  |  |  |  |
| 15-19 | 966 | 22.50 | 4.3 | 4.0 | 467.98<br>(p=0.000) |
| 20-25 | 3327 | 77.50 | 5.8 | 6.0 |  |
| All | 4293 | 100 | 5.6 | 5.5 |  |

<sup>a</sup>Kaplan–Meier estimates; <sup>b</sup>log rank $\chi^2$ test

**Figure 1.**
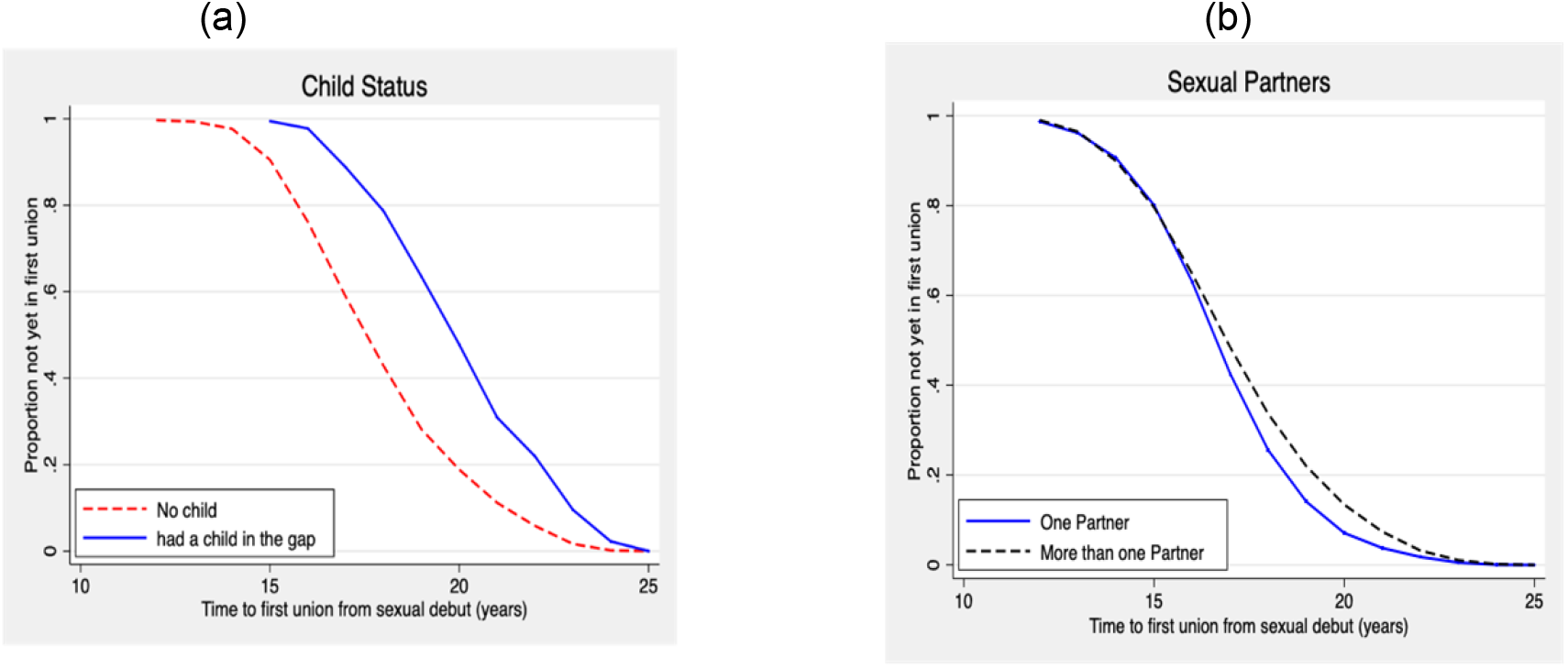
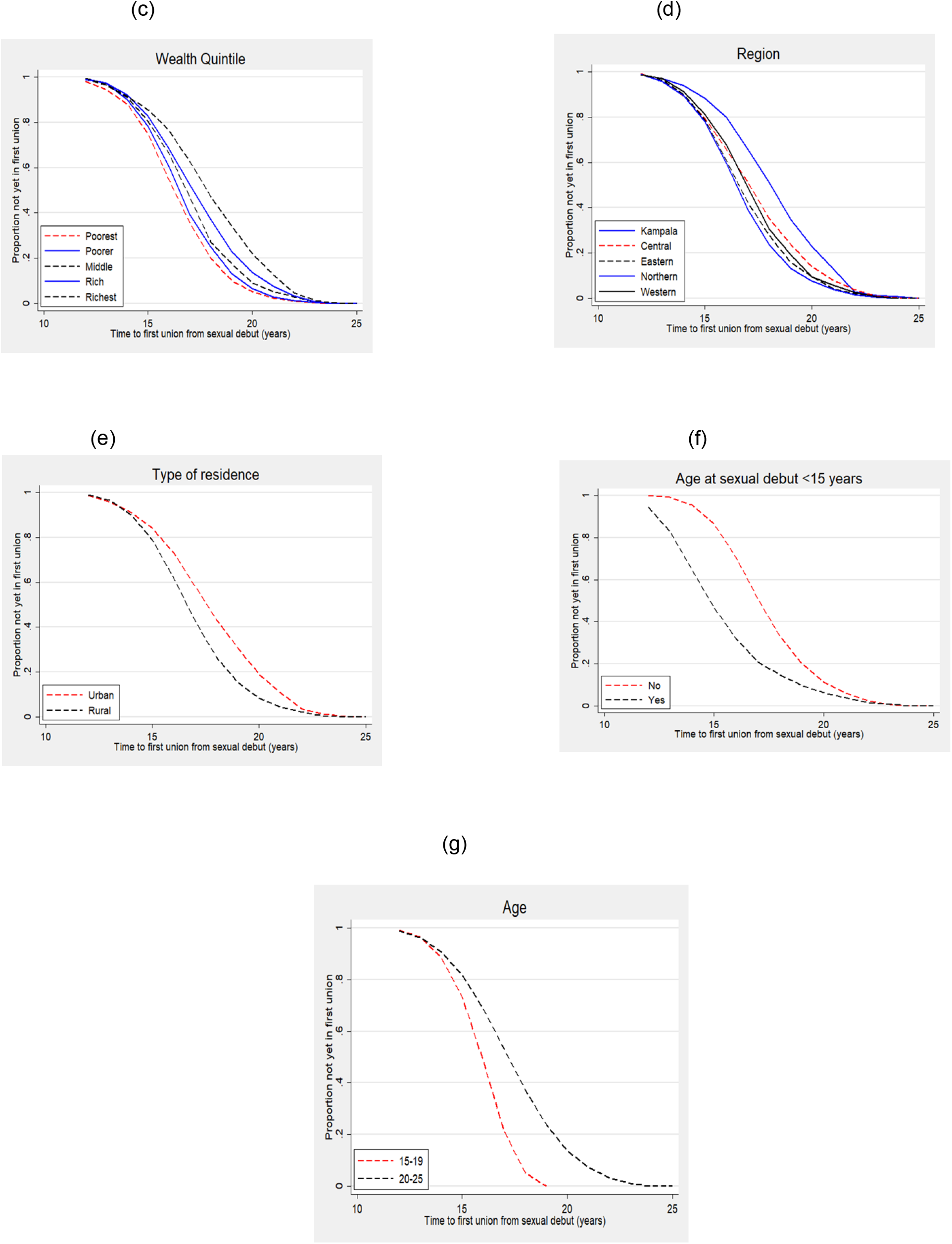
Kaplan-Meier plots showing the proportion of women who were not yet in their first union by selected direct, indirect and controlling factors: (a) Child Status, (b) Sexual partners, (c) Wealth quintile, (d) Region, (e) Type of residence, (f) age at sexual debut<15 years, and (g) current age of the woman.

With regard to the relationship between the timing of childbearing and the length of the gap, the table shows that about 23% of the women who had a birth in the gap prior to first union have a significantly longer gap (slightly above 8 years); and this is compared to nearly 77% of the women without a child whose transition to first union took nearly 6 years. The mean length of the gap increased monotonically with increase in the number of sexual partners, with women who have more than one partner, having a higher mean length of the interval of about 6 years; and those with one partner having a gap of nearly 5 years. Table 1 & Figure 1, demonstrates that this difference is strongly (χ^2^ =35.79; p=0.000). The mean length of the gap increases monotonically with each category of wealth index, women living in households with the wealthiest index having a mean gap of nearly 6 years compared to women who live in the poorest quintile whose length of the gap is about 5 years. Urban women have a long gap with a mean of 6.1 years compared to those who live in rural areas with a gap of about 5.3 years.

The regional distribution demonstrates that women who live in Kampala have longer intervals than those in Central, Eastern, Western or Northern with over 6 years of the gap between sexual debut and age at first union. Kampala region is followed by Central (5.7 years), Eastern (5.3 years), and Western (5.5 years). Women who live in the Northern region have the lowest mean length of the gap (5.2 years). The mean gap monotonically increases with woman’s age, with women in the age group 20-25 having the longest interval (5.8 years); and those in the youngest age group (15-19), having a mean length of the gap slightly above four years (4.3 years). As shown by Table 1 & Figure 1, a young age at sexual debut is associated with a significantly shorter interval (3.8 years). This variation signifies some historical differences in birth cohorts over historical time.

### Timing of first union after onset of sexual debut

Table 2 presents decrement lifetable estimates, in which, the results suggest that having a child in the gap between sexual debut and age at first marriage has a positive effect on women with regard to the marriage market. By the end of the fifth year, 63% of the women who had a child in the gap had entered first union compared to 76% of the women who were childless. The analysis show evidence about the observed difference (Wilcoxon-Gehan=85.37, p<0.000). This analysis underpins the view that having a premarital birth, especially on purpose, precipitates a quicker transition to first union. Having more than one sexual partner usually involves high risks especially in the era of HIV/AIDS. Though minimal, results in Table 2 suggest that there is a significant difference in the proportion of women transiting to first union (Wilcoxon-Gehan=17.45, p<0.000); with fewer women with multiple partners entering first union compared to women with one partner beyond the fifth year.

**Table 2.**
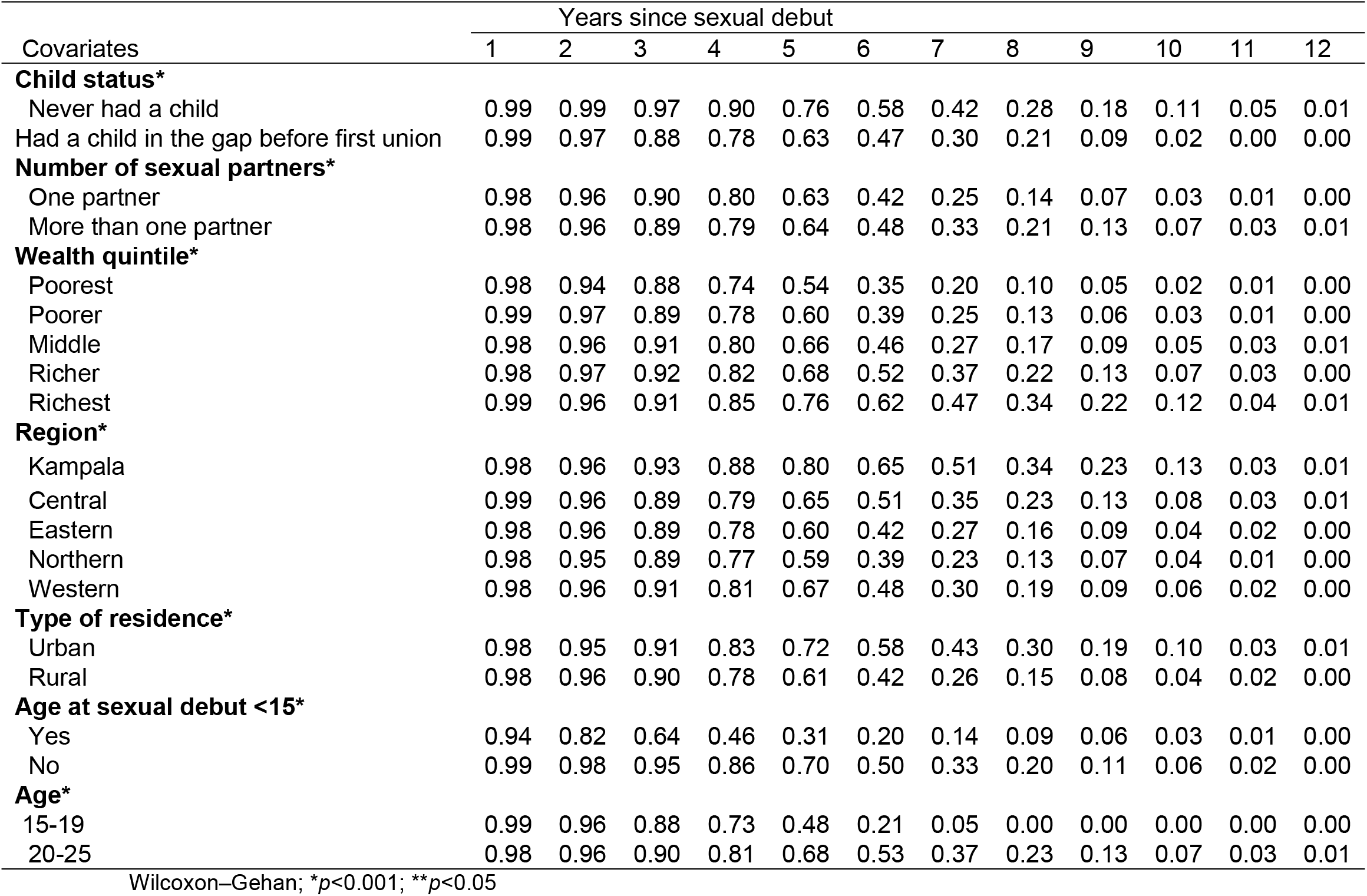
Decrement lifetable estimates showing the proportion of women who had their sexual debut but not yet in first union by some selected background characteristics, Uganda.

| Covariates | Years since sexual debut |  |  |  |  |  |  |  |  |  |  |  |
| --- | --- | --- | --- | --- | --- | --- | --- | --- | --- | --- | --- | --- |
|  | 1 | 2 | 3 | 4 | 5 | 6 | 7 | 8 | 9 | 10 | 11 | 12 |
| **Child status\*** |  |  |  |  |  |  |  |  |  |  |  |  |
| Never had a child | 0.99 | 0.99 | 0.97 | 0.90 | 0.76 | 0.58 | 0.42 | 0.28 | 0.18 | 0.11 | 0.05 | 0.01 |
| Had a child in the gap before first union | 0.99 | 0.97 | 0.88 | 0.78 | 0.63 | 0.47 | 0.30 | 0.21 | 0.09 | 0.02 | 0.00 | 0.00 |
| **Number of sexual partners\*** |  |  |  |  |  |  |  |  |  |  |  |  |
| One partner | 0.98 | 0.96 | 0.90 | 0.80 | 0.63 | 0.42 | 0.25 | 0.14 | 0.07 | 0.03 | 0.01 | 0.00 |
| More than one partner | 0.98 | 0.96 | 0.89 | 0.79 | 0.64 | 0.48 | 0.33 | 0.21 | 0.13 | 0.07 | 0.03 | 0.01 |
| **Wealth quintile\*** |  |  |  |  |  |  |  |  |  |  |  |  |
| Poorest | 0.98 | 0.94 | 0.88 | 0.74 | 0.54 | 0.35 | 0.20 | 0.10 | 0.05 | 0.02 | 0.01 | 0.00 |
| Poorer | 0.99 | 0.97 | 0.89 | 0.78 | 0.60 | 0.39 | 0.25 | 0.13 | 0.06 | 0.03 | 0.01 | 0.00 |
| Middle | 0.98 | 0.96 | 0.91 | 0.80 | 0.66 | 0.46 | 0.27 | 0.17 | 0.09 | 0.05 | 0.03 | 0.01 |
| Richer | 0.98 | 0.97 | 0.92 | 0.82 | 0.68 | 0.52 | 0.37 | 0.22 | 0.13 | 0.07 | 0.03 | 0.00 |
| Richest | 0.99 | 0.96 | 0.91 | 0.85 | 0.76 | 0.62 | 0.47 | 0.34 | 0.22 | 0.12 | 0.04 | 0.01 |
| **Region\*** |  |  |  |  |  |  |  |  |  |  |  |  |
| Kampala | 0.98 | 0.96 | 0.93 | 0.88 | 0.80 | 0.65 | 0.51 | 0.34 | 0.23 | 0.13 | 0.03 | 0.01 |
| Central | 0.99 | 0.96 | 0.89 | 0.79 | 0.65 | 0.51 | 0.35 | 0.23 | 0.13 | 0.08 | 0.03 | 0.01 |
| Eastern | 0.98 | 0.96 | 0.89 | 0.78 | 0.60 | 0.42 | 0.27 | 0.16 | 0.09 | 0.04 | 0.02 | 0.00 |
| Northern | 0.98 | 0.95 | 0.89 | 0.77 | 0.59 | 0.39 | 0.23 | 0.13 | 0.07 | 0.04 | 0.01 | 0.00 |
| Western | 0.98 | 0.96 | 0.91 | 0.81 | 0.67 | 0.48 | 0.30 | 0.19 | 0.09 | 0.06 | 0.02 | 0.00 |
| **Type of residence\*** |  |  |  |  |  |  |  |  |  |  |  |  |
| Urban | 0.98 | 0.95 | 0.91 | 0.83 | 0.72 | 0.58 | 0.43 | 0.30 | 0.19 | 0.10 | 0.03 | 0.01 |
| Rural | 0.98 | 0.96 | 0.90 | 0.78 | 0.61 | 0.42 | 0.26 | 0.15 | 0.08 | 0.04 | 0.02 | 0.00 |
| **Age at sexual debut <15\*** |  |  |  |  |  |  |  |  |  |  |  |  |
| Yes | 0.94 | 0.82 | 0.64 | 0.46 | 0.31 | 0.20 | 0.14 | 0.09 | 0.06 | 0.03 | 0.01 | 0.00 |
| No | 0.99 | 0.98 | 0.95 | 0.86 | 0.70 | 0.50 | 0.33 | 0.20 | 0.11 | 0.06 | 0.02 | 0.00 |
| **Age\*** |  |  |  |  |  |  |  |  |  |  |  |  |
| 15-19 | 0.99 | 0.96 | 0.88 | 0.73 | 0.48 | 0.21 | 0.05 | 0.00 | 0.00 | 0.00 | 0.00 | 0.00 |
| 20-25 | 0.98 | 0.96 | 0.90 | 0.81 | 0.68 | 0.53 | 0.37 | 0.23 | 0.13 | 0.07 | 0.03 | 0.01 |

Wilcoxon–Gehan; \* $p < 0.001$ ; \*\* $p < 0.05$

The transition to first union by region shows more women in the Northern (41%) to have entered first union by the fifth year after onset of sex initiation. The corresponding figures for Eastern, Western, Central and Kampala regions are 40%, 33%, 35% and 20%. Overall, a smaller proportion of women residing in Kampala or in other urban regions compared to women in other regions entered first union by the fifth year; and this variation is statistically significant (Wilcoxon-Gehan=78.3, p<0.000). With respect to the type of residence, the transition to first union was faster for women in rural areas compared to those in urban areas. A significant difference of 11% between rural and urban areas is observed (Wilcoxon-Gehan=80.83, p<0.000).

Table 2 also reveals significant variations by wealth quintile. Most women living in poorer households joined first union earlier than women than women residing in relatively wealth households. By the end of the fifth year, the proportions are; poorest (46%), poorer (40%), middle (34%), richer (32%) and richest (24%). Consistent with the mean length of the gap observed in Table 1 and Figure 1, Table 2 shows that the proportion of women entering first union is decreasing significantly and monotonically with increase in age (or age group). By the end of the fifth year, the proportions are: age group 15-19 (52%) and age group 20-25 (32%) and were significant (Wilcoxon-Gehan=331.67, p<0.000). Controlling for the early age at sexual debut (less than 15 years), the findings in Table 2 suggests that, for this sample, the proportion of women who entered first union following age at first sex before age 15 was higher compared to those whose sexual debut occurred at 15 years and older. In 5 years following sexual debut, 69% of such women had entered first union compared to 30%, and by the sixth year, a significant difference of 30% between those less than 15 and those aged 15 or older was observed (Wilcoxon-Gehan=588.69, p<0.000).

Table 3 presents the findings from the three discrete-time to event model analyses, conducted to examine the influence of some socioeconomic characteristics on the time taken to first union following sexual debut. Model I tested the effect of child status, number of sexual partners and wealth, on the odds that a respondent would report entry into first union. The effect of the presence of the child was negative but significant while the existence of a number of sexual partners was positive and significant. Nevertheless, there was no relationship with household wealth. In contrast to child status, sexual partners and wealth in Model I, Model II, which considers the effect of region of and type of residence on progress to first union, finds a negatively insignificant effect with number of sexual partners. Nevertheless, the effect remained associated with decreased risk of the presence of the child in the gap when other variables were entered into the model. The relationship between entry in first union and the age at sexual debut was positive and significant while that of the current age of the respondent was negative (Model III). Respondents whose sexual debut was above age 15 or older had higher odds of joining first union while those whose current age was 20-25 having reduced odds. The diagnostic test of the model suggests that the observed data reflected the expected (*hat:p=0*.*342; _hatsq:p= 0*.*269*).

**Table 3.**
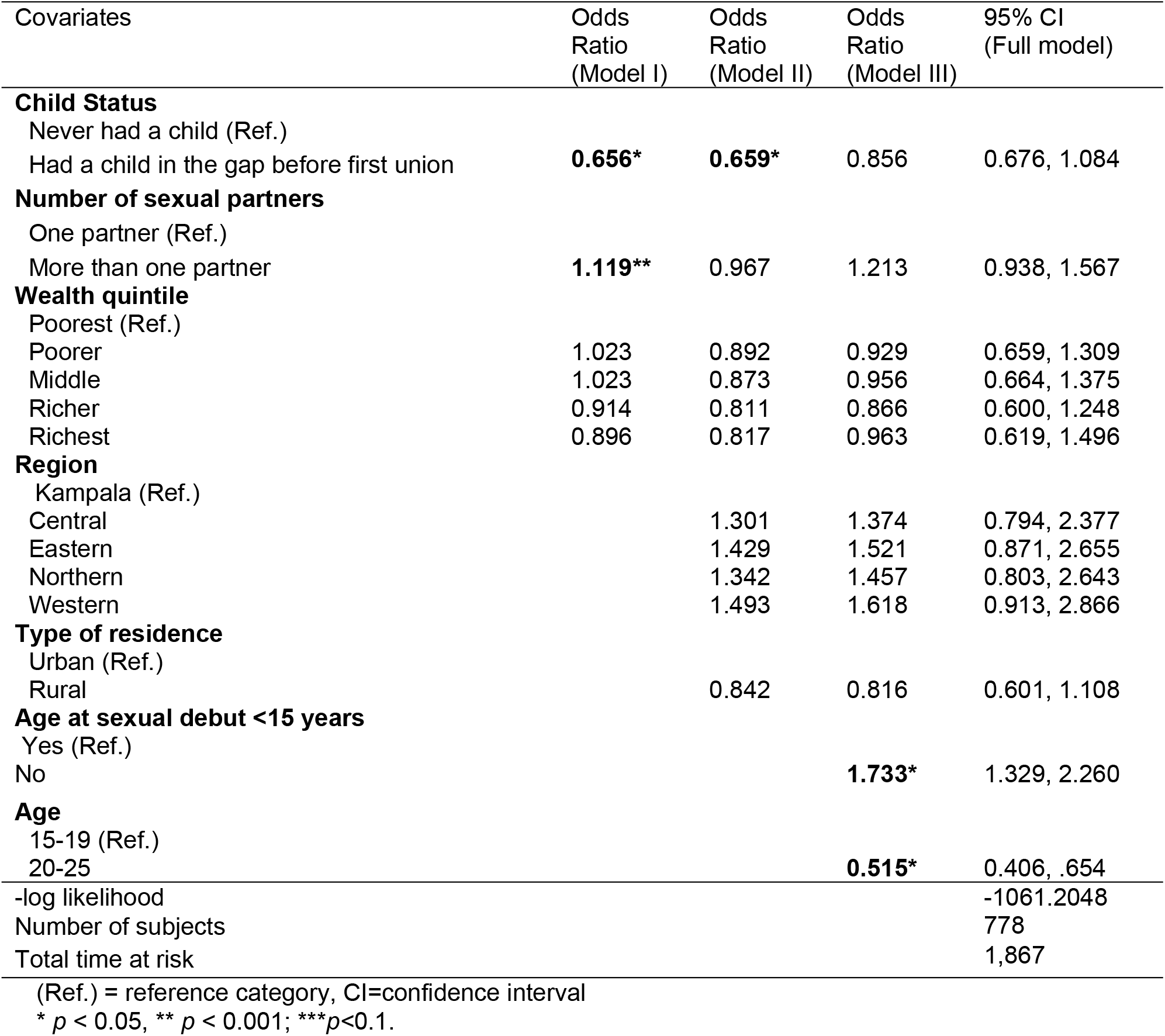
Results of the Discrete time logit model showing the relative risk of time to first union after sexual debut by some selected background characteristics among women aged 15–25,Uganda.

| Covariates | Odds Ratio (Model I) | Odds Ratio (Model II) | Odds Ratio (Model III) | 95% CI (Full model) |
| --- | --- | --- | --- | --- |
| **Child Status** |  |  |  |  |
| Never had a child (Ref.) |  |  |  |  |
| Had a child in the gap before first union | **0.656\*** | **0.659\*** | 0.856 | 0.676, 1.084 |
| **Number of sexual partners** |  |  |  |  |
| One partner (Ref.) |  |  |  |  |
| More than one partner | **1.119\*\*** | 0.967 | 1.213 | 0.938, 1.567 |
| **Wealth quintile** |  |  |  |  |
| Poorest (Ref.) |  |  |  |  |
| Poorer | 1.023 | 0.892 | 0.929 | 0.659, 1.309 |
| Middle | 1.023 | 0.873 | 0.956 | 0.664, 1.375 |
| Richer | 0.914 | 0.811 | 0.866 | 0.600, 1.248 |
| Richest | 0.896 | 0.817 | 0.963 | 0.619, 1.496 |
| **Region** |  |  |  |  |
| Kampala (Ref.) |  |  |  |  |
| Central |  | 1.301 | 1.374 | 0.794, 2.377 |
| Eastern |  | 1.429 | 1.521 | 0.871, 2.655 |
| Northern |  | 1.342 | 1.457 | 0.803, 2.643 |
| Western |  | 1.493 | 1.618 | 0.913, 2.866 |
| **Type of residence** |  |  |  |  |
| Urban (Ref.) |  |  |  |  |
| Rural |  | 0.842 | 0.816 | 0.601, 1.108 |
| **Age at sexual debut <15 years** |  |  |  |  |
| Yes (Ref.) |  |  |  |  |
| No |  |  | **1.733\*** | 1.329, 2.260 |
| **Age** |  |  |  |  |
| 15-19 (Ref.) |  |  |  |  |
| 20-25 |  |  | **0.515\*** | 0.406, .654 |
| -log likelihood |  |  |  | -1061.2048 |
| Number of subjects |  |  |  | 778 |
| Total time at risk |  |  |  | 1,867 |

(Ref.) = reference category, CI=confidence interval
\* $p < 0.05$ , \*\* $p < 0.001$ ; \*\*\* $p < 0.1$ .

#### Risk factors associated with the gap between sexual debut and age at first union

Table 3 presents results from the three logistic regression analyses we estimated to examine the correlates of the timing of first union among women in Uganda given sexual debut. In model I, we tested three variables including child status, number of sexual partners and wealth quintile on the odds that the respondent would report first union and found a strong link with two variables. Surprisingly, the effect of the number of sexual partners ceased to be significant when other variables (region and type of residence) were entered into the model (Model II). Indeed, in the full model (model III), only two effects (sexual debut and current age) are found to be significant. The effect of age at sexual debut as measured by whether it occurred before age 15 or 15 or over is significant and positive, and that of the current age measured whether the woman’s current age is 15-19 or 20-25, though significant, is negative. Regarding the diagnostic test of the full model, the link-test results show that the time to discrete model was well specified, with *hat: p=0*.*342 & _hatsq: p=0*.*269*, which implies that the observed data is almost the same as the expected data.

## Discussion

Despite the increasing empirical research on the age at sexual debut over the few decades, available studies have often limited their investigation on the woman’s reproductive health consequences, HIV infection and, trends in age at first sex. Yet, the age at first sex initiation, entails much more than just looking at an early age. This study has sought to fill this gap by adopting a novel focus on examining how sociodemographic factors associated with the length of the gap between onset of sex initiation and the age at first union later in life; estimating the waiting time to first union following sexual debut; and identifying the risk factors associated with the waiting time to first union. The descriptive findings show evidence of a short waiting time to first union following sexual debut. This finding is in line with those of Bozon et al who found similar results in their study in France (Bozon et al., 2012). This could be attributed to patriarchal beliefs, community expectation about girls in becoming wives and mothers, poverty, and limited opportunities for education resulting in fewer alternative opportunities. However, the waiting time is higher than that of Zaba and her colleagues finding from the Tanzanian population (Zaba et al., 2009).

While some socioeconomic characteristics appeared to be significantly associated with the length of the gap between sexual initiation and first union at bivariate level, only two factors– age at sexual debut and current age appear to play an important role in influencing this gap. Women whose age at sexual debut was between 15 and 25 were significantly more likely to transit to first union faster compared to women whose sexual initiation came earlier than age 15. Findings from previous studies about the empirical association between age at sexual debut and timing of first union in sub-Sahara Africa are scanty. However, delays in transiting to first union among women whose sexual debut came earlier than age 15 could be explained by two arguments: First, the practice of serial monogamy which is the habit of swiftly moving from one romantic relationship to another common among the young females(Yaya & Bishwajit, 2018); and second, the practice of having multiple and concurrent partners among young women which leads to a failure in subsequent union formation (Magnusson et al., 2015).

This study highlighted the role played by some demographic characteristics of women. The finding shows a negative link between the length of the gap and the woman’s current age. Women aged 20-25 currently had a disproportionally lower risk of transiting to first union within the sample following sexual initiation. In the Ugandan context, this could be attributed to social development aspects which result in a set of conditions that greatly diverge from earlier thinking that a woman is supposed to either be one’s wife or one’s mother. Two potential limitation of this study which could be addressed in future studies are: First, sex initiation and first union formation are complex constructs which were examined using self-reported data which may be subject to social desirability bias. Second, the Uganda Demographic Healthy Survey is cross-sectional, implying that what is reported are factors associated with, do not necessarily imply a causative relationship. Nonetheless, the study provides an important contribution to the inadequate extant literature on timing of first union following sexual debut. In conclusion, the study found the mean and median length of the gap between first union following sexual debut to be 5.6 and 5.5 years respectively.

Age at sexual debut and current age of the women were found to influence length of the gap between sexual initiation and first union.

## Data Availability

Acknowledgments. The authors extend their sincere gratitude to ICF International website (URL: https://www.dhsprogram.com/data/available-datasets.cfm) for allowing them to access the 2016 Uganda Demographic and Heathy Survey (UDHS) data.

## Acknowledgments

The authors extend their sincere gratitude to ICF International website (URL: https://www.dhsprogram.com/data/available-datasets.cfm) for allowing them to access the 2016 Uganda Demographic and Heathy Survey (UDHS) data.

## Funding

This research received no specific grant from any funding agency, commercial entity or not-for-profit organization. Nonetheless, the primary author is grateful to management of the College of Business and Management Sciences (CoBAMS), Makerere University, for the institutional grant, as research facilitation support, for staff.

## Conflicts of Interest

The authors have no conflicts of interest to declare.

## Ethical Approval

This study used secondary data and the authors declare that all procedures contributing to this work conform with the ethical standards of the relevant national and institutional committees on human experimentation and with the Helsinki Declaration of 1975, as revised in 2008.

## Notes

### Competing Interest Statement

The authors have declared no competing interest.

### Author Declarations

Ethical Approval. This study used secondary data and the authors declare that all procedures contributing to this work conform with the ethical standards of the relevant national and institutional committees on human experimentation and with the Helsinki Declaration of 1975, as revised in 2008.

